# N95 Filtering Facepiece Respirators Remain Effective After Extensive Reuse During the COVID-19 Pandemic

**DOI:** 10.1101/2020.12.04.20244087

**Authors:** Valeria Fabre, Sara E. Cosgrove, Yea-Jen Hsu, George Jones, Taylor Helsel, James Bukowski, Mark Sobota, Anna C. Sick-Samuels, Aaron M. Milstone, Lisa L. Maragakis, Clare Rock, for the Centers for Disease Control and Prevention Epicenters Program

## Abstract

In a cross-sectional evaluation of healthcare worker reuse of their own 3M N95 respirators, 83% (76/92) passed the seal check and the fit-test after a median of 40 repeated donnings. The user seal-check had 31% sensitivity to detect N95 respirator failure but 100% specificity.

## Background

The SARS-CoV-2 (COVID-19) pandemic has led to a widespread critical shortage of N95 respirators^1,2^. One conservation strategy is having healthcare workers (HCWs) reuse their own N95. Data on the safe number of N95 reuses before filtration failure is lacking. Based on a study in a simulated environment, the Centers for Disease Control and Prevention (CDC) suggests limiting the number of reuses to five per N95 to ensure an adequate safety margin^3^; however, such approach likely leads to discard of clinically effective N95s earlier than necessary^4^. Our primary objective was to evaluate effectiveness of reused N95s in a real-world healthcare setting during the COVID-19 pandemic. A secondary objective was to identify factors that could be used to proactively identify N95 failure.

## Methods

### Study setting

The Johns Hopkins Hospital (JHH), a 1,162-bed academic hospital in Baltimore, MD, recommends N95 respirators for any interaction with confirmed or suspected SARS-CoV-2 infected patients and when performing aerosol generating procedures for all patients, regardless of SARS-CoV-2 status. To preserve N95 supplies HCWs reuse their own N95s (covered with a face shield) until it must be discarded due to concern for compromised structural or functional integrity, as determined by visual inspection and a user performed seal check before every donning.

### Study design, participants, and definitions

We conducted a cross-sectional evaluation of reused N95 effectiveness during July and August 2020 in clinical areas dedicated for care of patients with known or suspected COVID-19: the Emergency Department and five inpatient units. On the day of the N95 assessment, participating HCWs were asked questions about reuse of their N95s included number of shifts worked with the current N95, average number of donnings per shift, longest number of hours and method for storing the N95. Question were answered based on best HCWs’ recall. Following the questionnaire, each N95 had a 3-step effectiveness screening: first, an inspection for structural damage (i.e., non-intact nosepiece or head straps, visible soilage), second, a user seal test to assess for air leakage during inhalation and exhalation, third a qualitative fit-testing with standard Saccharin Solution Aerosol Protocol performed by the study team^5^. Any N95 that failed the seal check or the saccharine fit-test was further evaluated with a confirmatory quantitative fit-test using the ambient aerosol condensation nuclei counter (PortaCount®) protocol^6,7^. A fit factor result on this protocol of < 100 is considered a failure. Due to constraints on N95 supply we used the quantitative method to evaluate only those N95s that failed a screening test as the PortaCount test requires insertion of a probe through the N95 to count particles rendering the N95 unusable. At the time of the study, the only available N95 models at our institution were 3M 1860 (dome-shape) and 3M 1870 (duck-bill). HCWs who failed the fit-test were given a new N95. The study was approved by JHM IRB.

### Statistical analysis

The primary outcome was a confirmed N95 failure, defined as failure of one or more screening tests followed by failure of the quantitative fit-test. Secondary outcomes included factors associated with failure, accuracy of the user seal check in detecting fit-failures. Fisher’s exact and Wilcoxon-rank sum tests were used to evaluate categorical and continuous variables. The relationship between number of repeated N95 donnings and N95 failure was assessed by Kaplan-Meier curves where survival was considered N95 passing. We conducted a sensitivity analysis including the N95s that failed a screening test (seal check or saccharin fit-test) but did not have a confirmatory PortaCount fit-test as failures. Based on data from preliminary observations, with 95% confidence interval and an error margin of two the required sample size was 77 HCWs. A 2-sided *P* value <0.05 was considered statistically significant. Analyses were performed using StataCorp 2019 (College Station, TX: StataCorp LLC).

## Results

Of 99 recruited HCWs, 92 had complete follow up (**Figure**) and were included in the primary analysis. The overall median number of self-reported N95 donnings at the time of the assessment of these 92 HCWs was 40 (IQR 17 – 100) and the median for the reported longest number of hours that the N95 was kept on once donned was 2.5 hours (IQR 1 – 2.5). All N95s (n=92) were structurally intact upon visual inspection, 80% (74/92) passed both the seal check and the saccharine fit-test, while 2 HCWs passed the seal check, failed the saccharine but passed the PortaCount fit-test (**Figure**), resulting in an overall pass rate of 83% (76/92) and a primary outcome of N95 failure in 17% (16/92) of N95s. Physicians and advanced practitioners were more likely to pass the fit-test (95%) compared to other roles (81% of bedside nurses, 66% technicians, *P*<0.01), **Table**. There was no difference by 3M N95 type, number of donnings, or reported folding of the N95 during storage, between N95s that did and did not pass the assessment.

**Table:** Participant characteristics and frequency of repeated N95 use by single healthcare workers.

| **Cohort characteristics** | **All**<br>**N=92** | **N95 pass**<br>**N= 76** | **N95 failure**<br>**N=16** | **P value** |
| --- | --- | --- | --- | --- |
| Gender, n (%) |  |  |  | 0.45 |
| Female | 77 (84) | 69 (82) | 15 (94) |  |
| Male | 15 (16) | 14 (18) | 1 (6) |  |
| Role, n (%) |  |  |  | 0.01 |
| Physician/Advanced practitioner | 22 (24) | 21 (28) | 1 (6) |  |
| Nurse | 54 (59) | 44 (58) | 10 (63) |  |
| Technician | 10 (11) | 5 (7) | 5 (31) |  |
| Other | 6 (6) | 6 (8) | 0 |  |
| Mask type, n (%) |  |  |  | 1.00 |
| 3M 1860 | 72 (78%) | 59 (78) | 13 (81) |  |
| 3M 1870 | 20 (22) | 17 (22) | 3 (19) |  |
| **N95 use** |  |  |  |  |
| Fold N95 for storage, n (%) |  |  |  | 0.47 |
| No | 62 (67) | 50 (66) | 12 (75) |  |
| Yes | 30 (33) | 26 (34) | 4 (25) |  |
| Visual Inspection, n (%) |  |  |  | 0.65 |
| Not creased | 83 (90) | 69 (91) | 10 (62) |  |
| Creased | 9 (10) | 7 (9) | 6 (38) |  |
| Duration of N95 reuse, n (%) |  |  |  | 0.11 |
| < 1 week | 7 (8) | 7 (9) | 0 |  |
| 1-4 weeks | 33 (36) | 24 (32) | 9 (56) |  |
| >4 weeks | 52 (56) | 45 (59) | 7 (44) |  |
| N95 donnings, median (IQR) | 40 (17.5-100) | 50 (20-120) | 40 (16 -82) | 0.39 |
| Longest hours N95 worn once<br>donned, median (IQR) | 2.5 (1-2.5) | 2.5 (1-2.5) | 2.5 (1-3.5) | 0.43 |

IQR: interquartile range.

**Figure:**
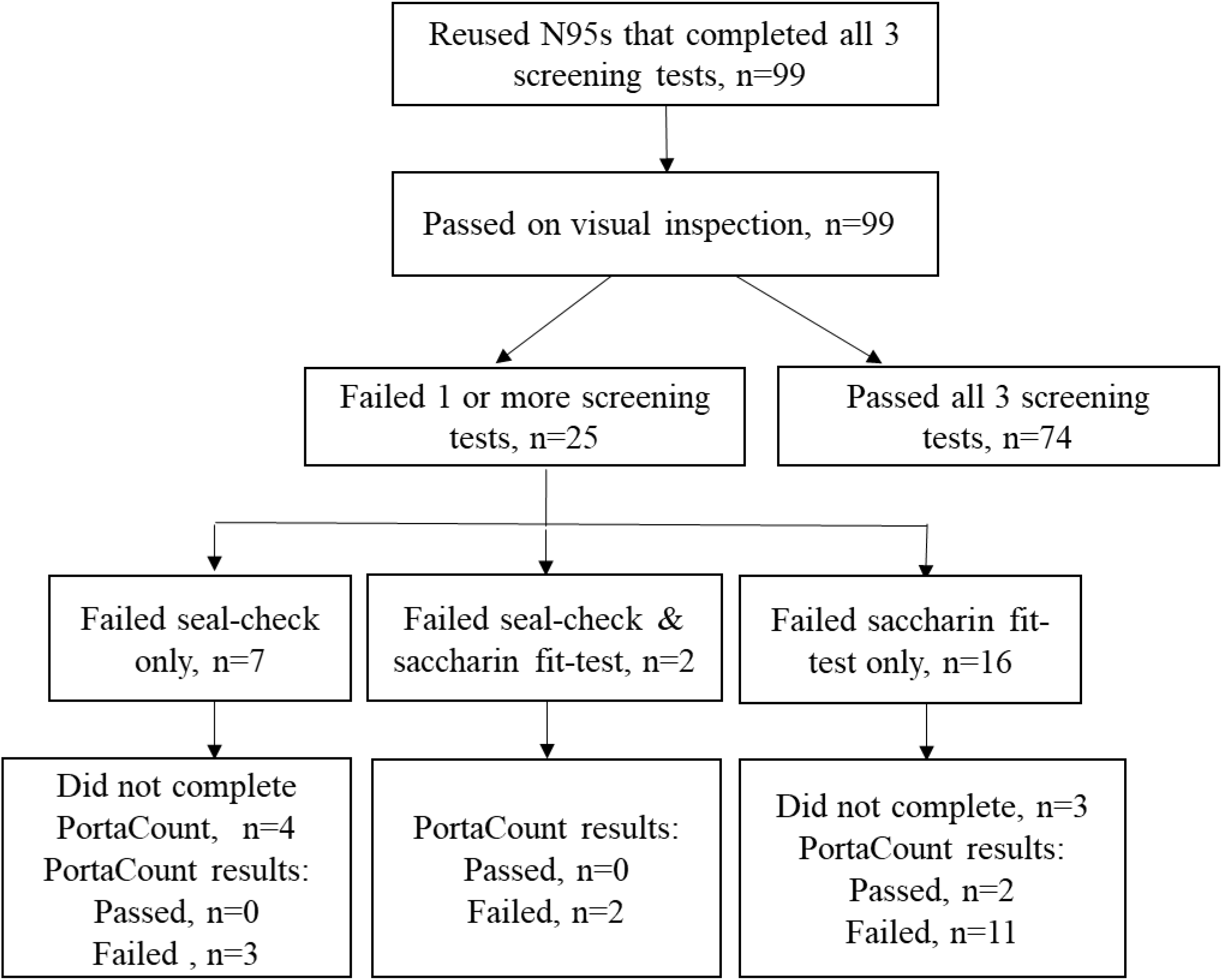
Flow diagram of recruited healthcare workers who reused N95s during the COVID-19 pandemic.

All N95s donned fewer than 12 times passed, and the probability of N95s maintaining a good fit was >95% for up to 23 donnings and >90% for up to 31 donnings (**Supplementary Figure 1 and Supplementary Table 1**).

The sensitivity of the user seal check to detect N95 fit-failure was 31% (5/16) and the specificity was 100% (5/5).

Cohort characteristics and N95 use factors associated with N95 failure remained similar in a sensitivity analysis that included 7 HCWs who failed a screening test and lacked confirmatory PortaCount data as failures (**Supplementary Table 2**). One HCW failed the seal check after 6 donnings. Like in the primary analysis, there were no saccharin failures before 12 donnings among HCWs who had passed a seal check and the probability of the N95 passing a fit check was >95% up to 16 repeated donnings and >90% up to 23 donnings (**Supplementary Table 3**).

## Discussion

This cross-sectional evaluation of HCWs reusing their own 3M N95s until visibly damaged, soiled or structurally unsound during the COVID-19 pandemic found that 83% were still effective as measured by fit-testing after a median of 40 reuses.

The user seal-check identified 31% of non-effective N95s with 100% specificity. This finding demonstrates the value of the user seal check, a simple, non-invasive fit check, recommended at each donning to identify gross leakage of air. The minimum reuse of N95s that passed the seal-check but failed the saccharin and PortaCount fit-test (covert N95 failures) was 12 times. Only certain HCW roles were associated with N95 failure, further evaluation including N95 reuse patterns would be needed to better understand this observation. We estimated the probability of N95s remaining effective at incremental N95 donnings and found that was >95% of N95 would maintain an adequate fit for up to 23 donnings and >90% for up to 31 donnings. Hence, these data suggest that, if critical N95 shortages exist, with an effective seal on donning, HCWs can safely reuse their N95s many times more than the number currently suggested by the CDC^3^.

Our study has limitations. We did not perform a PortaCount on N95s that passed the seal check or the saccharine fit-test due to limited N95 supplies and we may have overestimated “passes”; however, false passes are infrequent with the saccharin method^8^. Although we evaluated two of the most commonly used N95 respirators in the United States ^9^, findings may not be generalizable to alternative models. The number of repeated N95 donnings was based on HCW recall, which may have been under- or over-estimated; however, we do not think there was bias in either direction. We did not sample the N95s to assess for pathogen contamination, a risk of N95 reuse, but our protocol of face shield to protect N95 reduces this risk by preventing droplets from landing on the respirator surface. This study was not powered to assess effectiveness of N95s to prevent SARS-COV-2 infection or other potentially airborne transmitted infections. Notably, no patient-to-HCW SARS-COV-2 transmissions have been documented for HCWs who complied with the recommended COVID-19 precautions at JHH to date (authors’ personal communication). There was missing PortaCount data from some HCWs who failed the seal check or saccharine fit-test; however, we performed a sensitivity analysis to minimize the impact of missing data on interpretation of the study results.

In summary, extensive reuse of the N95 models tested in our study seems an acceptable and safe approach during critical supply shortages rather than uniform discarding of N95s after the currently suggested 5 reuses^3^ as long as HCWs consistently perform a seal check and obtain a good a seal before donning a reused N95 respirator. Consideration could be given to offering regular interval saccharine fit-testing when reusing N95s to enhance HCW comfort and safety with respirator reuse.

## Data Availability

All the data referred to in the manuscript is presented.

## Acknowledgements

We thank Alejandra B. Salinas for technical assistance and JHH HCWs for their participation in the study.

## Funding

Centers for Disease Control and Prevention Epicenter Program, COVID-19 supplement to grant 6 U01CK000554-02-02.

## SUPPLEMENTAL MATERIAL

**Supplementary Figure 1:**
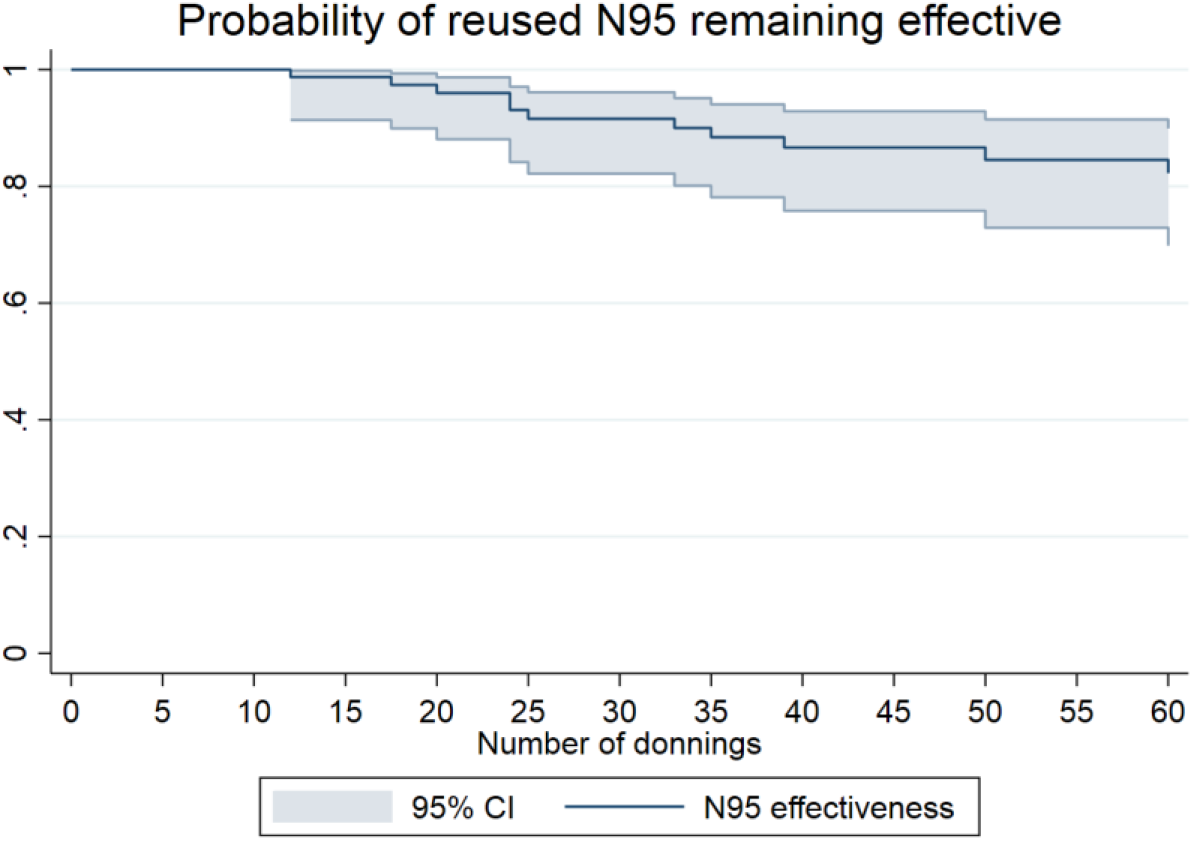
Probability of N95 maintaining a good fit after incremental donnings.

**Supplementary Table 1:** Probability of N95 maintaining a good fit with incremental donnings.

| Interval | Total | Failures per donning interval | Pass with no subsequent follow up | Passing | Error | 95% Confidence Interval |
| --- | --- | --- | --- | --- | --- | --- |
| 1 2 | 92 | 0 | 3 | 1 | 0 | . . |
| 3 4 | 89 | 0 | 1 | 1 | 0 | . . |
| 4 5 | 88 | 0 | 1 | 1 | 0 | . . |
| 5 6 | 87 | 0 | 1 | 1 | 0 | . . |
| 7 8 | 86 | 0 | 1 | 1 | 0 | . . |
| 10 11 | 85 | 0 | 6 | 1 | 0 | . . |
| 12 13 | 79 | 1 | 4 | 1 | 0.0129 | 0.9114 0.9982 |
| 15 16 | 74 | 0 | 1 | 0.9870 | 0.0129 | 0.9114 0.9982 |
| 17 18 | 73 | 1 | 2 | 0.9733 | 0.0186 | 0.8974 0.9933 |
| 20 21 | 70 | 1 | 2 | 0.9592 | 0.0231 | 0.8787 0.9867 |
| 22 23 | 67 | 0 | 1 | 0.9592 | 0.0231 | 0.8787 0.9867 |
| 24 25 | 66 | 2 | 2 | 0.9297 | 0.0304 | 0.8390 0.9702 |
| 25 26 | 62 | 1 | 0 | 0.9147 | 0.0334 | 0.8196 0.9608 |
| 28 29 | 61 | 0 | 1 | 0.9147 | 0.0334 | 0.8196 0.9608 |
| 30 31 | 60 | 0 | 2 | 0.9147 | 0.0334 | 0.8196 0.9608 |
| 33 34 | 58 | 1 | 0 | 0.8989 | 0.0363 | 0.7992 0.9506 |
| 35 36 | 57 | 1 | 2 | 0.8829 | 0.0391 | 0.7788 0.9398 |
| 36 37 | 54 | 0 | 3 | 0.8829 | 0.0391 | 0.7788 0.9398 |
| 37 38 | 51 | 0 | 1 | 0.8829 | 0.0391 | 0.7788 0.9398 |
| 39 40 | 50 | 1 | 0 | 0.8652 | 0.0421 | 0.7560 0.9278 |
| 40 41 | 49 | 0 | 2 | 0.8652 | 0.0421 | 0.7560 0.9278 |
| 42 43 | 47 | 0 | 1 | 0.8652 | 0.0421 | 0.7560 0.9278 |
| 45 46 | 46 | 0 | 3 | 0.8652 | 0.0421 | 0.7560 0.9278 |
| 48 49 | 43 | 0 | 1 | 0.8652 | 0.0421 | 0.7560 0.9278 |
| 49 50 | 42 | 0 | 1 | 0.8652 | 0.0421 | 0.7560 0.9278 |
| 50 51 | 41 | 1 | 2 | 0.8436 | 0.0463 | 0.7261 0.9136 |
| 52 53 | 38 | 0 | 1 | 0.8436 | 0.0463 | 0.7261 0.9136 |
| 60 61 | 37 | 1 | 1 | 0.8205 | 0.0504 | 0.6950 0.8980 |
| 63 64 | 35 | 0 | 2 | 0.8205 | 0.0504 | 0.6950 0.8980 |
| 66 67 | 33 | 0 | 1 | 0.8205 | 0.0504 | 0.6950 0.8980 |
| 70 71 | 32 | 0 | 2 | 0.8205 | 0.0504 | 0.6950 0.8980 |
| 73 74 | 30 | 0 | 1 | 0.8205 | 0.0504 | 0.6950 0.8980 |
| 75 76 | 29 | 0 | 1 | 0.8205 | 0.0504 | 0.6950 0.8980 |
| 80 81 | 28 | 0 | 2 | 0.8205 | 0.0504 | 0.6950 0.8980 |
| 84 85 | 26 | 0 | 1 | 0.8205 | 0.0504 | 0.6950 0.8980 |
| 100 101 | 25 | 1 | 0 | 0.7876 | 0.0581 | 0.6456 0.8779 |

**Supplementary Table 2.** Participant characteristics and fit pass rate on a sensitivity analysis where the seven healthcare workers who failed the seal check or the saccharin fit-test and had missing confirmatory PortaCount data are considered failures.

| Cohort characteristics | All, n=99 | Failure, n=23 | Pass, n = 76 | P value |
| --- | --- | --- | --- | --- |
| Gender, n (%) |  |  |  |  |
| Female | 82 | 20 (87) | 62 (82) | 0.54 |
| Male | 17 | 3 (13) | 14 (18) |  |
| Role, n (%) |  |  |  |  |
| Physician/Advanced practitioner | 24 | 3 (13) | 21 (28) | 0.10 |
| Nurse | 56 | 12 (53) | 44 (58) |  |
| Technician | 10 | 5 (50) | 5 (50) |  |
| Other | 9 | 3 (13) | 6 (8) |  |
| Mask type, n (%) |  |  |  |  |
| 3M 1860 | 74 | 15 (65) | 59 (78) | 0.23 |
| 3M 1870 | 25 | 8 (35) | 17 (22) |  |
| **N95 use characteristics** |  |  |  |  |
| Fold N95 for storage, n (%) |  |  |  |  |
| No | 68 | 12(18) | 56 (82) | 0.25 |
| Yes | 31 | 5 (22) | 26 (34) |  |
| User seal check, n (%) |  |  |  |  |
| Fail | 9 | 9 (39) | 0 | <0.01 |
| Pass | 90 | 14 (61) | 76 (100) |  |
| Duration N95 reuse, n (%) |  |  |  |  |
| < 1 week | 9 | 2 (9) | 7 (9) | 0.38 |
| 1-2 weeks | 18 | 7 (30) | 11 (14) |  |
| >2-4 weeks | 16 | 3 (13) | 13 (17) |  |
| > 4 weeks | 56 | 11 (45) | 45 (59) |  |
| Number of repeated donnings, median (IQR) | 43.5 (20-102) | 33 (17.5–60) | 45 (20–115) | 0.28 |
| Longest hours N95 worn after donning, median (IQR) | 2.5 (1-2.5) | 2.5 (1–2.5) | 2.5 (1–2.5) | 0.51 |

IQR: interquartile range.

**Supplementary Table 3:** Probability of passing the fit-test based on 23 failures (in this sensitivity analysis, the seven N95s that failed either the seal check or the saccharin test and that were not confirmed with PortaCount fit-test are considered failures).

| Interval | Total | Failures per donning interval | Pass with no subsequent follow up | Passing | Error | 95% Confidence Interval |
| --- | --- | --- | --- | --- | --- | --- |
| 1 2 | 99 | 0 | 3 | 1.0000 | 0.0000 | .. |
| 3 4 | 96 | 0 | 1 | 1.0000 | 0.0000 | .. |
| 4 5 | 95 | 0 | 1 | 1.0000 | 0.0000 | .. |
| 5 6 | 94 | 0 | 1 | 1.0000 | 0.0000 | .. |
| 7 8 | 93 | 1 | 1 | 0.9892 | 0.0108 | 0.9257 0.9985 |
| 10 11 | 91 | 0 | 6 | 0.9892 | 0.0108 | 0.9257 0.9985 |
| 11 12 | 85 | 1 | 0 | 0.9776 | 0.0157 | 0.9131 0.9943 |
| 12 13 | 84 | 1 | 4 | 0.9656 | 0.0195 | 0.8971 0.9888 |
| 14 15 | 79 | 1 | 0 | 0.9534 | 0.0228 | 0.8804 0.9823 |
| 15 16 | 78 | 0 | 1 | 0.9534 | 0.0228 | 0.8804 0.9823 |
| 17 18 | 77 | 2 | 2 | 0.9283 | 0.0283 | 0.8471 0.9672 |
| 20 21 | 73 | 1 | 2 | 0.9154 | 0.0307 | 0.8305 0.9588 |
| 21 22 | 70 | 1 | 0 | 0.9023 | 0.0329 | 0.8138 0.9500 |
| 22 23 | 69 | 0 | 1 | 0.9023 | 0.0329 | 0.8138 0.9500 |
| 24 25 | 68 | 2 | 2 | 0.8754 | 0.0370 | 0.7803 0.9311 |
| 25 26 | 64 | 1 | 0 | 0.8617 | 0.0389 | 0.7637 0.9212 |
| 28 29 | 63 | 0 | 1 | 0.8617 | 0.0389 | 0.7637 0.9212 |
| 30 31 | 62 | 0 | 2 | 0.8617 | 0.0389 | 0.7637 0.9212 |
| 33 34 | 60 | 1 | 0 | 0.8474 | 0.0408 | 0.7462 0.9106 |
| 35 36 | 59 | 1 | 2 | 0.8328 | 0.0426 | 0.7286 0.8996 |
| 36 37 | 56 | 0 | 3 | 0.8328 | 0.0426 | 0.7286 0.8996 |
| 37 38 | 53 | 0 | 1 | 0.8328 | 0.0426 | 0.7286 0.8996 |
| 39 40 | 52 | 1 | 0 | 0.8167 | 0.0447 | 0.7089 0.8877 |
| 40 41 | 51 | 0 | 2 | 0.8167 | 0.0447 | 0.7089 0.8877 |
| 42 43 | 49 | 0 | 1 | 0.8167 | 0.0447 | 0.7089 0.8877 |
| 45 46 | 48 | 0 | 3 | 0.8167 | 0.0447 | 0.7089 0.8877 |
| 48 49 | 45 | 0 | 1 | 0.8167 | 0.0447 | 0.7089 0.8877 |
| 49 50 | 44 | 0 | 1 | 0.8167 | 0.0447 | 0.7089 0.8877 |
| 50 51 | 43 | 2 | 2 | 0.7779 | 0.0503 | 0.6595 0.8593 |
| 52 53 | 39 | 0 | 1 | 0.7779 | 0.0503 | 0.6595 0.8593 |
| 56 57 | 38 | 1 | 0 | 0.7574 | 0.0530 | 0.6343 0.8440 |
| 60 61 | 37 | 1 | 1 | 0.7366 | 0.0555 | 0.6093 0.8281 |
| 63 64 | 35 | 0 | 2 | 0.7366 | 0.0555 | 0.6093 0.8281 |
| 66 67 | 33 | 0 | 1 | 0.7366 | 0.0555 | 0.6093 0.8281 |
| 70 71 | 32 | 0 | 2 | 0.7366 | 0.0555 | 0.6093 0.8281 |
| 73 74 | 30 | 0 | 1 | 0.7366 | 0.0555 | 0.6093 0.8281 |
| 75 76 | 29 | 0 | 1 | 0.7366 | 0.0555 | 0.6093 0.8281 |
| 80 81 | 28 | 0 | 2 | 0.7366 | 0.0555 | 0.6093 0.8281 |
| 84 85 | 26 | 0 | 1 | 0.7366 | 0.0555 | 0.6093 0.8281 |
| 100 101 | 25 | 1 | 0 | 0.7072 | 0.0606 | 0.5698 0.8078 |
| 105 106 | 24 | 0 | 1 | 0.7072 | 0.0606 | 0.5698 0.8078 |
| 115 116 | 23 | 0 | 1 | 0.7072 | 0.0606 | 0.5698 0.8078 |
| 120 121 | 22 | 1 | 2 | 0.6735 | 0.0664 | 0.5249 0.7847 |
| 122 123 | 19 | 1 | 1 | 0.6371 | 0.0721 | 0.4784 0.7591 |
| 133 134 | 17 | 0 | 1 | 0.6371 | 0.0721 | 0.4784 0.7591 |
| 156 157 | 16 | 0 | 1 | 0.6371 | 0.0721 | 0.4784 0.7591 |
| 162 163 | 15 | 0 | 1 | 0.6371 | 0.0721 | 0.4784 0.7591 |
| 180 181 | 14 | 1 | 1 | 0.5899 | 0.0807 | 0.4158 0.7280 |

## Notes

### Competing Interest Statement

The authors have declared no competing interest.

### Author Declarations

This study was approved by the Johns Hopkins Medicine IRB.

